# Limited impact of contact tracing in a University setting for COVID-19 due to asymptomatic transmission and social distancing

**DOI:** 10.1101/2021.11.10.21265739

**Authors:** Daniel Stocks, Emily Nixon, Adam Trickey, Martin Homer, Ellen Brooks-Pollock

## Abstract

Contact tracing is an important tool for controlling the spread of infectious diseases, including COVID-19. Here, we investigate the spread of COVID-19 and the effectiveness of contact tracing in a university population, using a data-driven ego-centric network model constructed with social contact data collected during 2020 and similar data collected in 2010. We find that during 2020, university staff and students consistently reported fewer social contacts than in 2010, however those contacts occurred more frequently and were of longer duration. We find that contact tracing in the presence of social distancing is less impactful than without social distancing. By combining multiple data sources, we show that University-aged populations are likely to develop asymptomatic COVID-19 infections. We find that asymptomatic index cases cannot be reliably back-traced through contact tracing and consequently transmission in their social network is not significantly reduced through contact tracing. In summary, social distancing restrictions had a large impact on limiting COVID-19 outbreaks in universities; to reduce transmission further contact tracing should be used in conjunction with alternative interventions.

## Introduction

Contact tracing is an important tool for controlling the spread of infectious diseases, including COVID-19. Contact tracing involves identifying individuals at high risk of infection due to their contact with a known case, and then testing, treating, or isolating them to prevent the onward spread of infection. It is commonly used for sexually transmitted infections [1, 37], where the definition of a contact is clearly established. Contact tracing is also used to control novel viral infections, particularly in the early containment phase of an epidemic [17, 23], as during the 2020/1 COVID-19 pandemic [18, 22].

The success of contact tracing depends on the proportion of transmission that occurs before symptom onset [15]; in the case of COVID-19, individuals are infectious for at least a day before developing symptoms and so *forward* contact tracing, which aims to identify contacts before they are infected, is likely to lag behind transmission. In contrast, *backward* contact tracing, which aims to identify missed ancestral cases and then use forward tracing to identify multiple branches of transmission, has been found to be more successful for COVID-19, in part due to the presence of super-spreaders [13].

Individual-based models are useful for capturing the heterogeneous nature of disease transmission and network models lend themselves to contact tracing analysis as they can explicitly define connections between individuals. We use an ego-centric network model framework, whereby a central completely connected node has alters that are satellites to the ego — a formulation that resembles an individual and their social contacts and can be constructed from available social contact data [11, 28].

The Social Contact Survey (SCS) collected data on social contact patterns for the general British population in 2010 and has been used for mathematical modelling of infectious disease [3, 10, 11]. However, university populations are different to the general population [12]. Students tend to be young adults, with higher than average numbers of social contacts and often live in communal residences. Also, during the COVID-19 pandemic, social restrictions have changed the way people socialise. To capture any new contact patterns in university staff and students during the pandemic, the Coronavirus Questionnaire (CON-QUEST) [28] survey was launched in June 2020.

With these two surveys we are able to investigate how social behaviours of a university population changed under social restrictions and then the effectiveness of contact tracing in this setting. The effectiveness of contact tracing is assessed by its ability to reduce the extent an individual spreads the disease and the schemes ability to back-trace and discover asymptomatic index cases.

## Methods

### Social Contact Data

#### The Social Contact Survey (SCS)

The SCS surveyed 5,388 individuals from across Great Britain in 2010; see [10, 11] for full details of the methodology and results. We extracted 635 individuals from the SCS whose occupation contained the text “Student”, “Lecturer” or “Researcher” and who were aged 18 or older. We assumed that these respondents were affiliated with a university.

#### The University of Bristol Coronavirus Questionnaire (CON-QUEST)

We used data from a longitudinal online survey of University of Bristol staff and students, launched on 23 June 2020. Here, we used data collected between 23 June 2020 and 8 February 2021. In that time, 744 staff and 887 students responded at least once and we used the first response from each participant. The CON-QUEST questionnaire was designed to mirror the SCS, so many of the questions are similar, but the surveys are not identical. In particular, SCS did not collect the age of the contact and CON-QUEST did not collect who-knows-whom data. For full details of CON-QUEST, see [28].

The data we extracted from CONQUEST was the age of the respondent, the age and number of individual contacts, the size of group contacts and the ages of their members, and the duration and frequency of any interaction. Note that ages of individual and group contacts were recorded within intervals, not as exact ages, so the ages of contacts are estimated by interpolating the intervals with a uniform distribution.

The duration and frequency distributions of social contacts reported in the two surveys are compared with a Chi-square test, to investigate whether social restrictions implemented during the COIVD-19 pandemic have changed contact patterns in the university.

### Modelling COVID-19 transmission and contact tracing on ego-centric networks

We model the spread of COVID-19 over ego-centric social networks, constructed from the social contact survey data described above. An ego-centric network consists of an ego, which is a central node that is connect to all other nodes, with alters that are connected to the ego but not necessarily to each other. This formulation lends itself to the available data when the respondent is modelled as the ego and their contacts are the alters. The edges represent social connections and are weighted with the duration (*T*) and frequency (*F*) of interaction. The nodes are weighted by the number of people involved in the interactions, so will only be greater than one 1 when representing a group.

Groups are assumed to be well-mixed and individuals group members are explicitly defined only during an interaction. Transmission within group is modelled on a virtual, complete network. The virtual network exists when a node weighted greater than 1 is interacted with. When the ego interacts with a group contact of size *s* (represented as a node with weight *s*) transmission is modelled on a complete network with *s* + 1 nodes and *s*(*s* + 1)*/*2 edges. How the respondent interacted with each group member is unknown, so we assume that the duration weighting of all edges in the group network is a fraction of the reported duration of the group interaction, such that the *s* edges of all nodes will be weighted with *T/s*, where *T* is the reported duration. Frequency weighting in the virtual network is unnecessary because the group interaction will already have been triggered. The edge that connects the ego with the weighted node will have a frequency weighting equal to the reported frequency.

We model contact tracing as originating from a symptomatic infectious node. Upon developing symptoms, symptomatic cases may be tested. All contacts they met in the past 5 days are traced. The symptomatic originating case and their traced contacts isolate. During isolation it is assumed that individuals contact no one, including household contacts. The only contacts that do not isolate are ones reported in CON-QUEST to be ‘met for the first time this day’, these contacts are considered strangers and unidentifiable.

### COVID-19 natural history parameters

We use a compartmental approach for capturing progression from susceptible to infection, to latent infection (not infectious), and infectious (either symptomatic or asymptomatic) [8, 31, 34].

We model the probability of transmission *P* (*T*) during an interaction to be dependent on the length of the interaction, *t*, as the risk of infection has been found to increase the longer a person is exposed to an infected individual [36]. We attempt to capture this time dependence of transmission with a stochastic transmission process that hinges on an adaptation of the fixed infectious period model reported in [19]. We consider asymptomatic individuals to be less infectious by a factor *A*; the probability of transmission during an interaction, *P* (*T*), is then

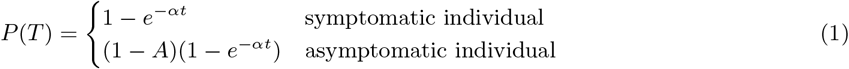

where *α* is the rate of transmission and *t* is the duration of interaction in minutes. We used *A* = 0.35 [7], and values of *α* that are: 1.6 *×* 10^−4^, 3.1 *×* 10^−4^, 5.6 *×* 10^−4^and 7.9 *×* 10^−4^ per minute. These values are based on data from the SCS, where the median number of unique contacts was 9, the median frequency of interaction is 6 days in two weeks and mean duration was 124 minutes. Taken together with an infectious period of 12.2 days, this corresponds to an average number of secondary cases between 1 and 4.

Upon infection, all individuals become latently infected with a mean latent period of 3.3 days [38]. Subsequently, the infectious period is modelled as lasting for 10 days after symptom onset, to reflect the UK track and trace guidelines which state that a person who has developed symptoms of COVID-19 must isolate for 10 days [27].

Infectious individuals are either symptomatic or asymptomatic. Age is a key factor in the probability of symptoms, with younger individuals less likely to report typical COVID-19-like symptoms [24, 35], so we seek to quantify an age-dependent probability of being asymptomatic. Measuring the age dependence of being asymptomatic directly is challenging, therefore we calculate the conditional probability using Bayes’ Theorem,

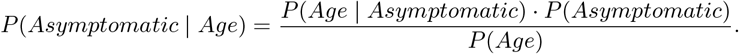

where *P* (*Age* | *Asymptomatic*) is the age distribution of asymptomatic cases, estimated using a meta-analysis [21], *P* (*Asymptomatic*) is the mean percentage of asymptomatic cases in infected individuals [29] and *P* (*Age*) is the probability of an individual belonging to a given an age group, calculated from data on the UK age demographic [14]. The banding of age groups as well as their conditional probability of being asymptomatic are shown in Table 1.

**Table 1:**
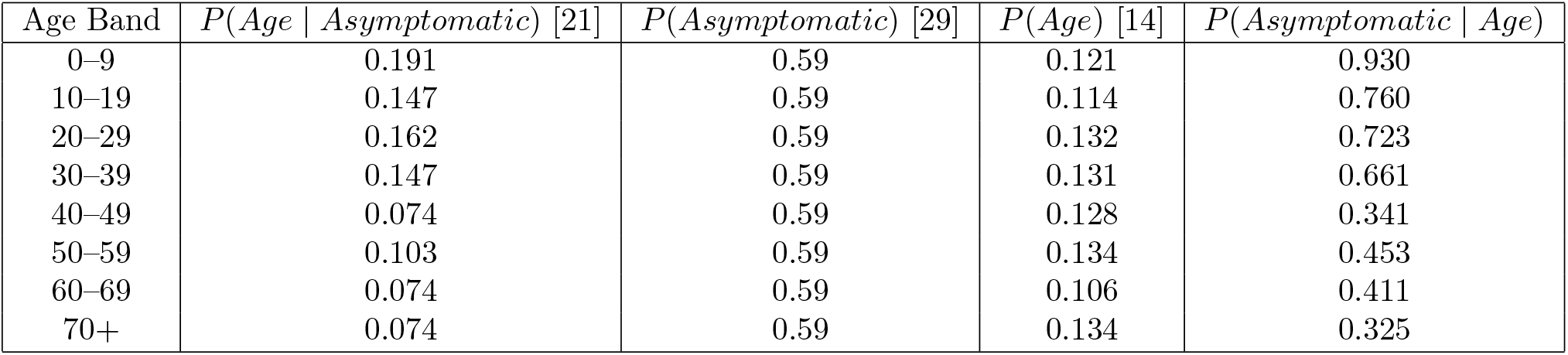
Conditional probabilities of an individual being asymptomatic given their age with the constituent parts of Bayes’ Theorem used to calculate the conditional probabilities. *P* (*Age* | *Asymptomatic*) is taken to be the proportion of asymptomatic cases that fell into each age band in [21]. *P* (*Asymptomatic*) here, is the mean proportion of cases found to be asymptomatic across the studies reviewed in [29]. *P* (*Age*) is the proportion of people in the UK that fall into the each age band, according to the 2017 ONS records on the demographic of the UK [14]. In the fifth column, which contains *P* (*Asymptomatic Age*), shows that in general the probability of being asymptomatic decreases with age. The only deviation is in the age band 40–49.

| Age Band | $P(\text{Age} \mid \text{Asymptomatic})$ [21] | $P(\text{Asymptomatic})$ [29] | $P(\text{Age})$ [14] | $P(\text{Asymptomatic} \mid \text{Age})$ |
| --- | --- | --- | --- | --- |
| 0–9 | 0.191 | 0.59 | 0.121 | 0.930 |
| 10–19 | 0.147 | 0.59 | 0.114 | 0.760 |
| 20–29 | 0.162 | 0.59 | 0.132 | 0.723 |
| 30–39 | 0.147 | 0.59 | 0.131 | 0.661 |
| 40–49 | 0.074 | 0.59 | 0.128 | 0.341 |
| 50–59 | 0.103 | 0.59 | 0.134 | 0.453 |
| 60–69 | 0.074 | 0.59 | 0.106 | 0.411 |
| 70+ | 0.074 | 0.59 | 0.134 | 0.325 |

### The Simulation Procedure

Event-based simulations with a time step of one day are used to model the local epidemic. The simulations are initiated with an infected ego and susceptible alters, and run for 30 days. On any given day, two nodes that are connected by an edge have a probability of interacting equal to *F/*14, where *F* is the frequency weighting (the expected number of interactions in 14 days) of the connecting edge. The probability of transmission during this interaction is given by equation 1. All newly infected individuals transition through the stages of a COVID-19 infection described above.

Alters in the network develop asymptomatic infections with a probability dependent on their age (Table 1). There are three scenarios for the egos symptomatic status: (1) they are all symptomatic, (2) they are all asymptomatic, they are asymptomatic with a probability dependent of their age (Table 1). Once an individual develops symptoms it is assumed they take a test that same day and receive the results within 3 days. Individuals will not isolate until they receive their result, and it at this point that all identifiable contacts met in the past 5 days also isolate. Isolation is implemented by setting the probability that an individual meets their contacts to zero. Isolation lasts for 10 days for the individual who tests positive and 14 days for their contacts. An individual is considered susceptible once their isolation ends. Hospitalisations and deaths are not included in this model as we are focusing on a primarily young population.

## Results

### Comparing characteristics between the two surveys

The median number of daily contacts reported in the CON-QUEST survey was 2, compared to 9 from the university-affiliated subset of the SCS used here. No one in the SCS subset reported having zero contacts on the previous day, but this was reported by 18% of CON-QUEST participants. The proportion of respondents with low numbers of daily contacts is greater in the CON-QUEST data than in the SCS data. The proportion of respondents reporting one, two or three daily contacts is an order of magnitude greater in the CON-QUEST survey than in the SCS subset (Figure 1d). The maximum number of daily contacts was greater in the SCS subset (*k* = 3011) than in CON-QUEST (*k* = 531) and there was a higher overall proportion of respondents with high numbers of contacts in the SCS subset (the proportion of respondents reporting 12–24 contacts was one order of magnitude greater).

**Figure 1:**
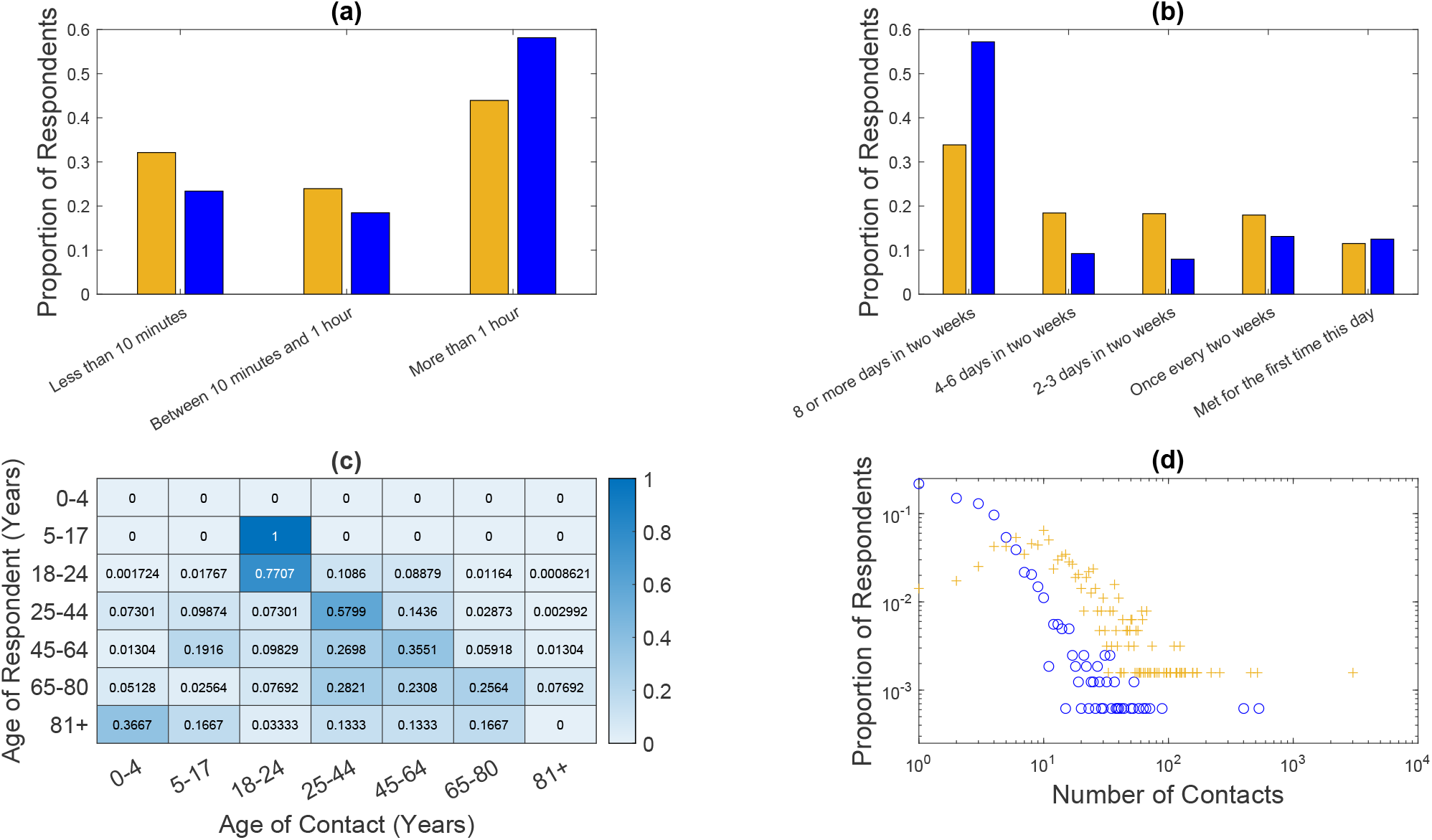
Comparison of social contact data for university students and staff in a pre-COVID-19 pandemic survey (the 2009 Social Contacts Survey, n = 635) [10, 11] and COroNavirus QUESTionnaire (June 2020 to February 2021, n = 1631) [28]. For all panels, data shown in blue in taken from the CON-QUEST and data in orange is taken from the SCS. **(a)** The interaction durations between the respondents and their contacts. **(b)** The interaction frequencies between the respondents and their contacts. **(c)** The proportion of contacts within each age category reported by respondents in each age category in CON-QUEST. **(d)** The distribution of the number of daily contacts (on a loglog scale). Both datasets are normalised by their own sizes

**Figure 2:**
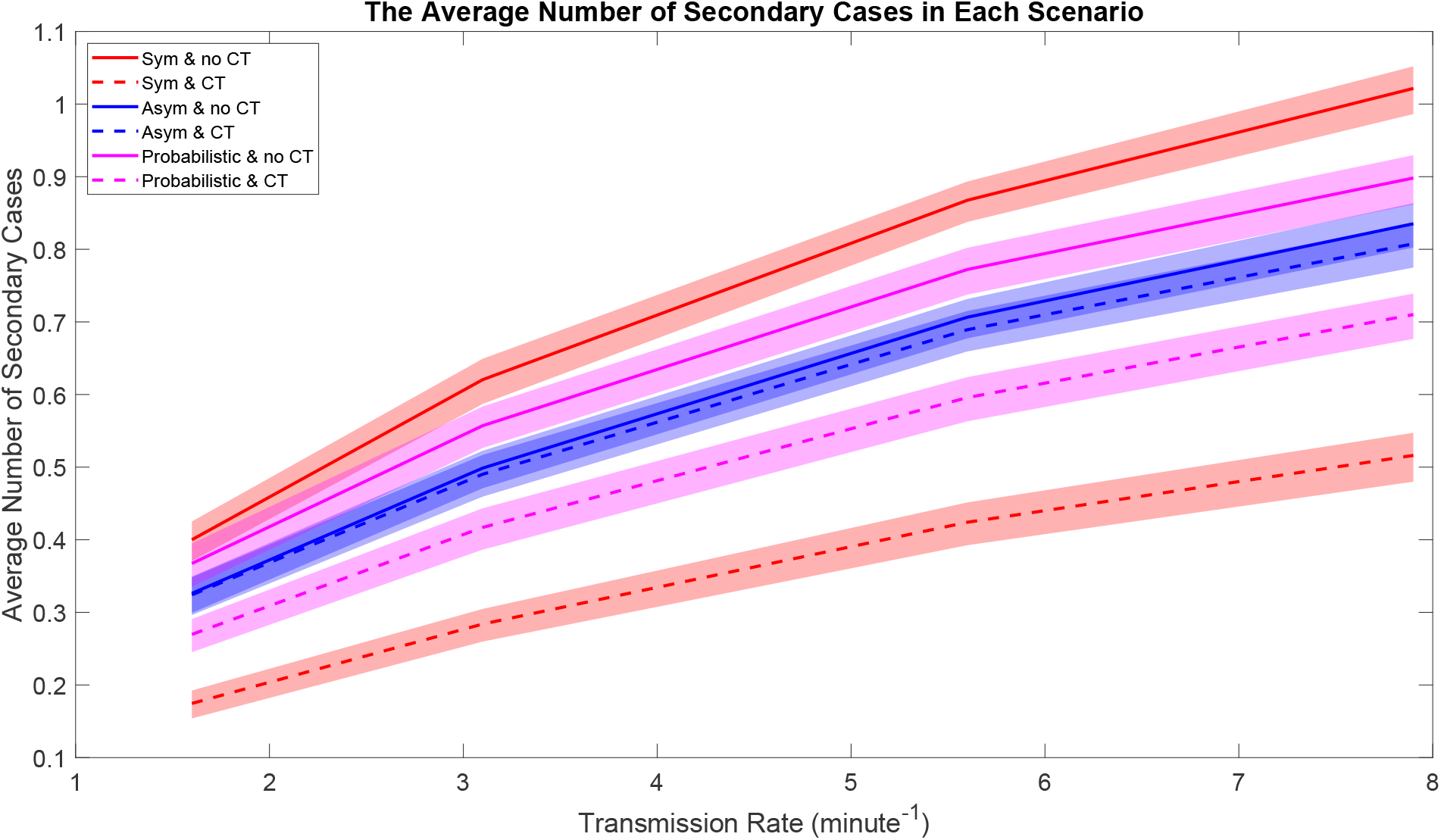
The average number of secondary cases in each scenario plotted against the transmission rate (*α*). The lines show the mean number of secondary cases per index case, in their respective scenarios and the coloured regions around each line show the 95% confidence intervals of the respective means.

The duration of social contacts reported during 2020 increased compared to contacts reported in 2010 (*χ*^2^ = 367.8, df = 2, *p* 0.001). Respondents in the CON-QUEST survey had longer interactions with their contacts than those in the SCS, with 58% and 43% of respondents having interactions that lasted longer than one hour respectively. A higher percentage of university-affiliated respondents in the SCS reported short interactions lasting of less than 10 minutes (33%) compared to the CON-QUEST survey (24%). In the SCS, nearly 60% of all reported interactions lasted less than one hour, compared to 42% in CON-QUEST (Figure 1a).

In addition to longer duration contacts, social contacts reported in CON-QUEST occurred more frequently than in the SCS (*χ*^2^ = 1518, df = 4, *p* 0.001). A greater proportion of contacts reported to be met ‘8 or more times in 2 weeks’ in the CON-QUEST survey than in the SCS (57.2% and 29.2% respectively). However, there was little difference between the number of contacts that were ‘Met for the first time this day’ (SCS-13.8%, CON-QUEST-12.5%) (Figure 1b).

For respondents with ages 18–24 (n = 637) and 25–44 (n = 647) the majority of their contacts (77% and 58% respectively) were within their own age interval. Respondents aged 45–64 (n = 314) had the greatest proportion of their contacts in their own age group (36%) but not a majority, while respondents 65–80 (n = 16) had similar proportions of contacts aged 25–44, 45–64 and 65–80 (28%, 23%, 25% respectively). However, respondents aged 81+ (n = 1), reported no contacts in their own age category. The greatest proportion of contacts for respondents age 81+ was 0–4 years old.

### Secondary COVID-19 cases

To assess how observed behavioural changes have impacted transmission and highlight the importance of contextually accurate data in modelling, the model is run with both university affiliated data from the SCS and the CON-QUEST data. In these simulations it is assumed the ego is symptomatic and there is no contact tracing. As can be seen in Table 2, when the ego-centric networks are constructed from the pre-COVID data recorded in the SCS there is a greater average number of secondary cases for all transmission rates. The average number of secondary cases is at least 1 when the SCS data is used. However, only with the highest transmission rate (fitted to produce a reproductive number of 4 in a simplified pre-COVID social network) does the average number of secondary cases reach 1 when the CON-QUEST data is used.

**Table 2:** The mean and 95% confidence intervals for average number of secondary infections when the simulation is run using data from the pre-COVID data from the SCS and the data from CON-QUEST. *α*_1*-*4_ are the transmission rates provided in Table 1.

| Transmission rate | SCS | CON-QUEST |
| --- | --- | --- |
| $\alpha_1$ | 1.1 [1.0, 1.2] | 0.40 [0.38, 0.42] |
| $\alpha_2$ | 1.9 [1.9, 2.0] | 0.62 [0.59, 0.65] |
| $\alpha_3$ | 2.7 [2.6, 2.8] | 0.87 [0.84, 0.89] |
| $\alpha_4$ | 3.2 [3.1, 3.3] | 1.0 [0.99, 1.0] |

### The effect of contact tracing

The spread of COVID-19 in a university population is found to be dependent on the transmission rate, the infectiousness of the index case, and whether contact tracing is implemented. Figure **??** shows that an increase in transmission rate always results in an increase in the average number of secondary cases. However, it is only in the scenario where all index cases are symptomatic and the transmission rate is 7.9 *×* 10^−4^ per minute that the average number of secondary cases exceeds 1 (Figure **??**a).

In the absence of contact tracing, symptomatic index cases generate a greater number of secondary cases compared to asymptomatic index cases due to their increased infectiousness. However, the spread of COVID-19 from symptomatic index cases is greatly reduced with the implementation of contact tracing. In comparison, the reduction due to contact tracing for asymptomatic index cases is minimal. When the index case is symptomatic, contact tracing has the potential to reduce the mean number of secondary cases by up to 56%, with the biggest impact for lower transmission rates. When the index case is asymptomatic, contact tracing reduces the mean number of secondary cases by less than 3%. In the asymptomatic case, the greater impact is seen with higher transmission probabilities. When the index case has a probability of being asymptomatic dependent on their age we see the expected behaviour of COVID-19 in a university population, shown in the bottom panel of Figure **??**. In this realistic scenario, contact tracing has the potential to reduce the mean number of secondary cases by 26.6%, 25.2%, 22.8% and 20.9% for *α*_1*-*4_ respectively.

The inability for contact tracing to curb asymptomatic transmission can be explained by the low percentage of index cases that are back-traced from their symptomatic contacts. As the transmission rate is increased in the model from *α*_1_ to *α*_4_ the percentage of index cases isolated also increases, from 4% to 12%, but remains low. This is inline with the increasing relative reduction of average cases seen in the asymptomatic scenario.

## Discussion

Analysis comparing the two sets of survey data suggests university populations have become more clustered during the pandemic. The general pattern of the CON-QUEST data is that university populations have few select contacts that they meet frequently for long periods of time. This pattern is in line with expected lockdown behaviour. The increase in clustering within university populations seen in the 2020 data meaningfully reduces the average number of secondary cases produced by an infectious individual. This reduction is seen when model simulations are run with the pre-COVID SCS data and the CON-Q UEST data. The comparison suggests that social restrictions can prevent COVID-19 from becoming endemic in a university for all but the most transmissible variants. However, there may be other explanations for the differences seen between the SCS and CON-QUEST surveys. For example, a survey in 2017/8, which collected information on contact patterns of UK citizens [20], found a real reduction in the number of contacts made by 15-19 year-olds from data collected 2005/6, that could be explained by the digitisation of young people’s social interactions [32]. Therefore, the differences seen in the social contact behaviour in students captured in the 2010 SCS compared to the behaviour captured by CONQUEST in 2020 may not be completely attributable to the COVID-19 pandemic.

When contact tracing is used in combination with social restrictions, they amount to an effective control. For all transmission rates when contact tracing is used the average number of secondary cases is below one. However, the extent to which contact tracing reduces transmission is limited when index cases are asymptomatic, suggesting that asymptomatic transmission cannot be curbed by contact tracing when asymptomatic cases are left undetected. Considering that universities have a young population and younger people have a higher probability of being asymptomatic, contact tracing may not sufficiently reduce transmission without mass testing. With mass testing the expected university behaviour will more closely resemble the scenario where all index cases are symptomatic.

The inability of contact tracing to control asymptomatic transmission partly stems from the fact that young members of the university interact most with other young members. Within these cliques an asymptomatic invective is likely to transmit to a person who will also develop an asymptomatic infection. When transmission is low, the majority of index cases will only transmit to people who develop asymptomatic infections. Hence, contact tracing cannot be relied upon to back-trace and isolate an asymptomatic index case.

These findings are consistent with other modelling approaches in identifying social distancing and mass testing as necessary measures for control in a university. Modelling conducted with anonymised university accommodation data found that asymptomatic cases produce more secondary cases even if they have a lower transmission rate than symptomatic cases [2]. A non-data driven model for COVID-19 transmission on US campuses also found that mass testing is important but does conclude that when prevalence is low, high rates of testing can lead to high proportions of isolated individuals with false-positives [30]. These high proportions of unnecessarily isolated people were generated without the inclusion of contact tracing; only those that were tested were isolated. With the low levels of transmission found in this model, if contacts identified through mass testing isolated then the number of unnecessary isolations would be a justifiable criticism of the control measure.

Although CON-QUEST is a longitudinal survey, we only analyse the first responses from the 23^rd^ June 2020 to 8^th^ February 2021. In this period, there was variation in the social restrictions measures and we do not capture such time varying behaviour here, nor do we consider significant social events such as mass travel for University vacations.

Our model does not account for vaccination or natural immunity and the results are representative of early stages in the pandemic, which is true also for the contact data used in constructing the networks. Other models have found that vaccination can reduce COVID-19 incidence in a population to near zero [25], even without other mitigating controls [4]. Due to the low levels of transmission observed in the simulations, we would expect few reinfections in the majority of networks. However, due to heterogeneity in the networks, where multiple transmissions do occur it is likely they will be to a select few individuals and here reinfection is a possibility. Consequently, the inclusion of natural immunity would likely lead to a reduction in the average number of secondary cases.

Further limitations of the model are that alters are not connected, and the assumption is made that social networks are isolated from each other, where in reality population level connections do exist. More extensive data collection such as that done by the SCS, where respondents reported which of their contacts had met each other, would provide information on transitive links. In the absence of these data, algorithms such as preferential attachment, an observed social phenomenon [5, 26], could be used. With regards to population-level networks, more targeted data collection could be conducted on schools or departments within universities [6, 9, 33], collecting information on whole cohort interactions. Data collection on this scale can be expensive and at times impractical, so synthetic social networks can be construct via algorithms. Previous research [16] has modelled COVID-19 transmission assuming a scale-free degree distribution exists at a population level, and investigated how different population-level social restrictions can reduce the critical levels for herd immunity. However, both the SCS and CON-QUEST data suggest that the degree distribution of a university population is power-law distributed only in the tail. Better population-level networks could be constructed through data-driven algorithms, such as exponential random graph models.

## Conclusion

In this paper we compare the results of two social surveys conducted before and during the pandemic, by looking at the frequency, duration and number of contacts that were reported. From the data we construct ego-centric networks to model COVID-19 transmission from an index cases to their contacts. We then introduce contact tracing and investigate three scenarios where the index case is either, always symptomatic, always asymptomatic, or asymptomatic with a probability dependent on their age.

During the period of active social restrictions the contact patterns of university populations changed, people generally met fewer contacts for longer periods and with a greater frequency. This behavioural change reduces the average number of secondary infections. Further reduction can be achieved with contact tracing, when cases are identifiable. In a university population where there is expected to be a large asymptomatic population, mass testing will be required to identify asymptomatic individuals and exercise contact tracing to its full potential. However, our model suggests that under social restrictions population level out-breaks are unlikely in a university.

## Data Availability

Data use in the present study are available request at: http://data.bris.ac.uk/data/dataset/2jxe5mx7gzbku2dekvlmbcwwhk or an anonymised version is available from: https://github.com/DStocks42/Uni-Contact-Tracing-Code. Other data used is openly accessible from: http://wrap.warwick.ac.uk/54273/

http://data.bris.ac.uk/data/dataset/2jxe5mx7gzbku2dekvlmbcwwhk

## Acknowledgements

EBP would like to acknowledge support from the National Institute for Health Research (NIHR) Health Protection Research Unit (HPRU) in Behavioural Science and Evaluation at the University of Bristol. The views expressed are those of the author(s) and not necessarily those of the NIHR or the Department of Health and Social Care. EBP, EN and DS are supported by UKRI through the JUNIPER consortium (Grant Number MR/V038613/1). EBP is further supported by MRC (Grant Number MC/PC/19067). AT is funded by the Wellcome Trust through his Sir Henry Wellcome Fellowship.

## Data availability

The CONQUEST data used in this study is available from Nixon, E. J. (Creator), Trickey, A. J. W. (Creator) & Brooks Pollock, E. (Creator), University of Bristol, 18 May 2021

DOI: 10.5523/bris.2jxe5mx7gzbku2dekvlmbcwwhk, http://data.bris.ac.uk/data/dataset/2jxe5mx7gzbku2dekvlmbcwwhk

The SCS data used in this study is available from Danon, L. (Creator), Read, J. M. (Creator), Keeling, M. J. (Creator), House, T. A. (Creator) and Vernon, M. C. (Creator), University of Warwick, 2009. http://wrap.warwick.ac.uk/54273/

## References

[1] Brewer, D., “Case-Finding Effectiveness of Partner Notification and Cluster Investigation for Sexually Transmitted Diseases/HIV”, in: Sex Transm Dis 32.2 (2005), pp. 78–83, doi: 10.1097/01.olq.0000153574.38764.0e.

[2] Brooks-Pollock, E., Christensen, H., Trickey, A., et al., “High COVID-19 transmission potential associated with re-opening universities can be mitigated with layered interventions”, in: Nat comm 12.12 (2021), p. 5017, doi: 10.1038/s41467-021-25169-3.

[3] Brooks-Pollock, E., Read, J. M., McLean, A. R., et al., “Mapping social distancing measures to the reproduction number for COVID-19”, in: Phil Trans R Soc B 376.1829 (2021), P20200276, doi: 10.1098/rstb.2020.0276.

[4] Bubar, K. M., Reinholt, K., Kissler, S. M., et al., “Model-informed COVID-19 vaccine prioritization strategies by age and serostatus”, in: Science 371.6532 (2021), pp. 916–921, doi: 10.1126/science.abe6959.

[5] Capocci, A., Servedio, V. D. P., Colaiori, F., et al., “Preferential attachment in the growth of social networks: The internet encyclopedia Wikipedia”, in: Phy Rev 74.3 (2006), p. 036116, doi: 10.1103/PhysRevE.74.036116.

[6] Cauchemez, S., Bhattarai, A., Marchbanks, T. L., et al., “Role of social networks in shaping disease transmission during a community outbreak of 2009 H1N1 pandemic influenza”, in: PNAS 108.7 (2011), pp. 2825–2830, doi: 10.1073/pnas.1008895108.

[7] Chen, Y., Wang, A., Yi, B., et al., “Epidemiological analysis of infection among close contacts of novel coronavirus pneumonia in Ningbo”, in: Chinese Journal of Epidemiology 41.5 (2020), pp. 667–671.

[8] Chung, N. N. and Chew, L. Y., “Modelling Singapore COVID-19 pandemic with a SEIR multiplex network model”, in: Sci Rep 11.1 (2021), p. 10122, doi: 10.1038/s41598-021-89515-7.

[9] Conlan, A. J. K., Eames, K. T. D., Gage, J. A., et al., “Measuring social networks in British primary schools through scientific engagement”, in: Proc. R. Soc. B. 278.1711 (2010), pp. 1467–1475, doi: 10.1098/rspb.2010.1807.

[10] Danon, L., Read, J. M., House, T. A., et al., “Social Encounter Networks: Collective Properties and Disease Transmission”, in: J.R.Soc.Interface 9.76 (2012), pp. 2826–2833, doi: 10.1098/rsif.2012.0357.

[11] Danon, L. et al., “Social encounter networks: characterizing Great Britain”, in: Proc Biol Sci 280.1765 (2013), doi: 10.1098/rspb.2013.1037.

[12] Elmer, T., Mepham, K., and Stadtfeld, C., “Comparisons of Students’ Social Networks and Mental Health Before and During the COVID-19 Crisis in Switzerland”, in: PLoS One 15.7 (2020), doi: 10.1371/journal.pone.0236337.

[13] Endo, A., Abbott, S., et al., “Estimating the overdispersion in COVID-19 transmission using outbreak sizes outside China”, in: Wellcome Open Res 5 (2020), p. 67, doi: 10.12688/wellcomeopenres.15842.3.

[14] Gov.uk, Overview of the UK population: July 2017, https://www.ons.gov.uk/peoplepopulationandcommunity/populationandmigration/populationestimates/articles/overviewoftheukpopulation/july2017, (accessed: 10/03/2021).

[15] Hellewell, J., Abbott, S., Gimma, A., et al., “Feasibility of controlling COVID-19 outbreaks by isolation of cases and contacts”, in: Lancet Glob Health 8.4 (2020), e488–e496, doi: 10.1016/S2214-109X(20)30074-7.

[16] Herman, H. A. and Schwartz, J.-M., “Why COVID-19 models should incorporate the network of social interactions”, in: Phys Bio 17.6 (2020), p. 065008, doi: 10.1088/1478-3975/aba8ec.

[17] Kang, M., Song, T., Zhong, H., et al., “Contact Tracing for Imported Case of Middle East Respiratory Syndrome, China, 2015”, in: Emerg Infect Dis 22.9 (2016), pp. 1644–6, doi: 10.3201/eid2209.152116.

[18] Keeling, M. J., Hollingsworth, T. D., and Read, J. M., “Efficacy of Contact Tracing for the Containment of the 2019 Novel Coronavirus (COVID-19)”, in: J Epidemiol Community Health 74.10 (2020), pp. 861–866, doi: 10.1136/jech-2020-214051.

[19] Keeling, M. J. and Grenfell, B. T., “Individual-based Perspectives on R0”, in: Journal of Theoretical Biology 203.1 (2000), pp. 51–61, doi: 10.1006/jtbi.1999.1064.

[20] Klepac, P., Kucharski, A. J., Conlan, A. J., et al., “Contacts in context: large-scale setting-specific social mixing matrices from the BBC Pandemic project”, in: 2020, doi: 10.1101/2020.02.16.20023754.

[21] Kronbichler, A. et al., “Asymptomatic patients as a source of COVID-19 infections: A systematic review and meta-analysis”, in: Int J Infect Dis 98 (2020), pp. 180–186, doi: 10.1016/j.ijid.2020.06.052.

[22] Kucharski, A. J. and al., et, “Effectiveness of isolation, testing, contact tracing, and physical distancing on reducing transmission of SARS-CoV-2 in different settings: a mathematical modelling study”, in: Lancet Infect Dis 20.10 (2020), pp. 1151–1160.

[23] Mclean, E., Pebody, R. G., Campbell, C., et al., “Pandemic (H1N1) 2009 influenza in the UK: clinical and epidemiological findings from the first few hundred (FF100) cases”, in: Epidemiol Infect 138.11 (2010), pp. 1531–41, doi: 0.1017/S0950268810001366.

[24] McQuade, E. T. R., Guertin, K. A., Becker, L., et al., “Assessment of Seroprevalence of SARS-CoV-2 and Risk Factors Associated With COVID-19 Infection Among Outpatients in Virginia.”, in: JAMA Network Open 4.2 (2021), e2035234, doi: 0.1001/jamanetworkopen.2020.35234.

[25] Moghadas, S. M., Vilches, T. N., Zhang, K., et al., “The Impact of Vaccination on Coronavirus Disease 2019 (COVID-19) Outbreaks in the United States”, in: Clinical Infectious Diseases (2021), ciab079, doi: 10.1093/cid/ciab079.

[26] Newman, M. E. J., “Clustering and preferential attachment in growing networks”, in: Phys Rev 64.2 (), 025102(R), doi: https://doi.org/10.1103/PhysRevE.64.025102.

[27] NHS, When to self-isolate and what to do, https://www.nhs.uk/conditions/coronavirus-covid-19/selfisolation-and-treatment/when-to-self-isolate-and-what-to-do/, (accessed: 23/03/2021).

[28] Nixon, E., Trickey, A., Christensen, H., et al., “Contacts and behaviours of university students during the COVID-19 pandemic at the start of the 2020/21 academic year”, in: Sci Rep 11 (2021), doi: 10.1038/s41598-021-91156-9.

[29] Oran, D. and Topol, E., “Prevalence of Asymptomatic SARS-CoV-2 Infection : A Narrative Review”, in: Ann Intern Med 173.5 (2020), pp. 362–367, doi: 10.7326/M20-3012.

[30] Paltiel, A. D., Zheng, A., and Walensky, R. P., “Assessment of SARS-CoV-2 Screening Strategies to Permit the Safe Reopening of College Campuses in the United States”, in: JAMA Netw Open 3.7 (2020), e2016818, doi: 10.1001/jamanetworkopen.2020.16818.

[31] Radŭlescu, A., Williams, C., and Cavanagh, K., “Management strategies in an SEIR-type model of COVID 19 community spread”, in: Sci Rep 10.1 (2020), p. 21256, doi: 10.1038/s41598-020-77628-4.

[32] Rideout, V. and Robb, M. B., Social medial, social life: teens reveal their experiences. Technical Report, https://www.commonsensemedia.org/sites/default/files/uploads/research/2018_cs_socialmediasociallife_fullreport-final-release_2_lowres.pdf, 2018.

[33] Salathé, M., Kazandjieva, M., Lee, J. W., et al., “A high-resolution human contact network for infectious disease transmission”, in: PNAS 107.51 (2010), pp. 22020–22025, doi: 10.1073/pnas.1009094108.

[34] Teslya, A., Pham, T. M., Godijk, N. G., et al., “Impact of self-imposed prevention measures and short-term government-imposed social distancing on mitigating and delaying a COVID-19 epidemic: A modelling study”, in: PLoS 17.7 (2020), e1003166, doi: 10.1371/journal.pmed.1003166.

[35] Wells, P. M., Doores, K. J., Couvreur, S., et al., “Estimates of the rate of infection and asymptomatic COVID-19 disease in a population sample from SE England”, in: J. Infect 81.6 (2020), pp. 931–936, doi: 10.1016/j.jinf.2020.10.011.

[36] Wiersinga, W. J., Rhodes, A., Cheng, A. C., et al., “Pathophysiology, Transmission, Diagnosis, and Treatment of Coronavirus Disease 2019 (COVID-19) A Review”, in: JAMA 324.8 (2020), pp. 782–793, doi: 10.1001/jama.2020.12839.

[37] Willcox, R., Jefferiss, F., and Naughten, E., “Contact Investigation of male West Indian Patients with Gonorrhoea”, in: British Journal of Venereal Diseases 42.3 (1966), p. 167.

[38] Zhao, S., “Estimating the time interval between transmission generations when negative values occur in the serial interval data: using COVID-19 as an example”, in: MBE 17.4 (2020), doi: 10.3934/mbe.2020198.

